# Plasma phosphorylated tau 181 and 217 as biomarkers for multiple sclerosis diagnosis, subtyping, and prognosis

**DOI:** 10.1101/2025.04.30.25326752

**Authors:** Chen Hu, Xuemei Zeng, Lili Zhang, Anuradha Sehrawat, Megan Powell, Emily Song, Elizabeth LS Walker, Alexis Watterson, Wen Zhu, Thomas K Karikari, Zongqi Xia

## Abstract

**Background:** Blood-based biomarkers are crucial for individualized management of multiple sclerosis (MS). Blood neurofilament light chain (NfL) and glial fibrillary acidic protein (GFAP) have shown promising clinical utility in MS, but they are insufficient to guide clinical management. Plasma tau proteins remain underexplored despite the growing evidence of shared pathology in Alzheimer’s disease and MS. We aimed to: (1) assess the utility of plasma tau biomarkers (phosphorylated tau 181 [p-tau181], p-tau217, and total tau [t-tau]) in MS diagnosis, subtyping, and prognosis; and (2) compare their performance with NfL and GFAP.

**Methods:** From a clinic-based prospective cohort, we included 160 people with MS (pwMS; 117 with relapsing-remitting MS [RRMS], 43 with progressive MS [PMS]) and 20 non-MS controls, all with baseline plasma samples. We measured baseline plasma concentrations of p-tau181, p-tau217, t-tau, NfL, and GFAP using ultrasensitive immunoassays. We collected demographics, clinical information, and longitudinal multi-modal outcomes (Patient Determined Disease Steps [PDDS], normalized age-related MS severity score [ARMSS], walking speed, manual dexterity, cognitive performance, retinal nerve fiber layer [RNFL] thickness, total brain volume, and gray matter volume) over a median follow-up of 3.0 years (IQR, 3.5). Adjusting for demographic and clinical covariates, we evaluated associations between biomarkers and MS diagnosis, subtypes, and prognosis. We examined the enhanced value of tau markers, in addition to NfL and GFAP, for subtype distinction and outcome prediction. Participants were enrolled between 2017 and 2023. Analyses were conducted in December 2024.

**Results:** Participants (n=180) had a median age of 51 years and were predominantly women (68%) and non-Hispanic white (91%). Compared with controls, pwMS had higher levels of p-tau217 (1.0 vs 0.7 pg/mL; p=0.04) and NfL (14.1 vs 9.0 pg/mL; p<0.01). Among pwMS, higher p-tau181 (aOR [95%CI]=2.3 [1.4,4.1]) and p-tau217 (aOR [95%CI]=3.0 [1.8,5.7]) were associated with PMS. These markers improved MS subtype classification accuracy beyond clinical features, NfL, and GFAP. Higher baseline p-tau181 and p-tau217 predicted worse disability, functional outcomes, and imaging outcomes independent of other biomarkers.

**Conclusions:** Plasma p-tau181 and p-tau217 are promising biomarkers for MS subtype classification and disability prediction, providing complementary information to NfL and GFAP. Further studies to validate their potential clinical utility in guiding MS management are warranted.

## Introduction

Multiple sclerosis (MS) is a chronic disease of the central nervous system (CNS) causing progressive accumulation of neurological disability.^1,2^ Based on clinical course, MS subtypes include relapsing–remitting (RRMS), primary progressive (PPMS), or secondary progressive (SPMS), and inform disease-modifying therapy (DMT) selection.^3^ Complex pathophysiological mechanisms drive variable disease progression trajectories among people with multiple sclerosis (pwMS).^4,5^ Fluid biomarkers offer promise to guide individualized management.^6–8^ Cerebrospinal fluid (CSF) biomarkers have limited clinical feasibility.^9–11^ In contrast, blood biomarkers are less invasive to collect, more reproducible to assay, and better suited for longitudinal monitoring in chronic neurological disorders. Recent advances in assay development have expanded the availability and reliability of blood-based biomarkers, providing opportunities to investigate their potential utility in MS.^12,13^

Blood neurofilament light chain (NfL) and glial fibrillary acidic protein (GFAP) disease have shown promise to inform MS disease activity and progression.^14–18^ However, their capabilities to diagnose MS and differentiate MS subtypes are limited.^19^ In current practice, MS subtypes are retrospectively confirmed using clinical history and magnetic resonance imaging (MRI). This limitation delays the recognition of patients with progressive subtypes and hinders timely clinical decisions (*e.g.,* given the limited DMT options for PMS). Thus, the field needs accessible biomarkers that distinguish RRMS from PMS early in MS course.^20^

Tau protein forms, well-known biomarkers of neurodegenerative diseases, may complement NfL and GFAP in MS.^21–25^ Previous studies reported tau accumulation in CSF of pwMS relative to non-inflammatory neurological controls.^26,27,28^ Abnormal tau phosphorylation was observed in brain tissue of pwMS and experimental autoimmune encephalomyelitis models, suggesting tau pathology as relevant in MS.^29–31^ Notably, tau phosphorylation in the CSF of PPMS was higher than RRMS.^32^ To our knowledge, there has been no report of blood tau biomarkers for MS.

Here, we measured plasma total tau (t-tau), phosphorylated tau 181 (p-tau181), phosphorylated tau 217 (p-tau217), NfL and GFAP by leveraging the biobank of a clinic-based prospective MS cohort. We examined whether plasma tau proteins provide additional clinical utility beyond existing biomarkers (*i.e.,* NfL, GFAP) in MS diagnosis, subtype differentiation, and prognosis (of long-term multi-modal outcomes).

## Methods

### Ethics approval

The University of Pittsburgh Institutional Review Board approved the study protocols (STUDY19080007). All participants provided written informed consent.

### Study population

Participants enrolled in the Prospective Investigation of Multiple Sclerosis in the Three Rivers Region (PROMOTE), a clinic-based prospective cohort (UPMC, Pittsburgh) that has enrolled adults ≥18 years since 2017. For this study, we defined pwMS as having a neurologist-confirmed MS diagnosis according to the 2017 McDonald criteria.^33^ We defined participants without a diagnosis of MS or related disorders (*i.e.*, neuromyelitis optica spectrum disorder, myelin oligodendrocyte glycoprotein antibody-associated disease, and other neuroimmunological disorders) as controls (**Figure 1**).

**Figure 1.**
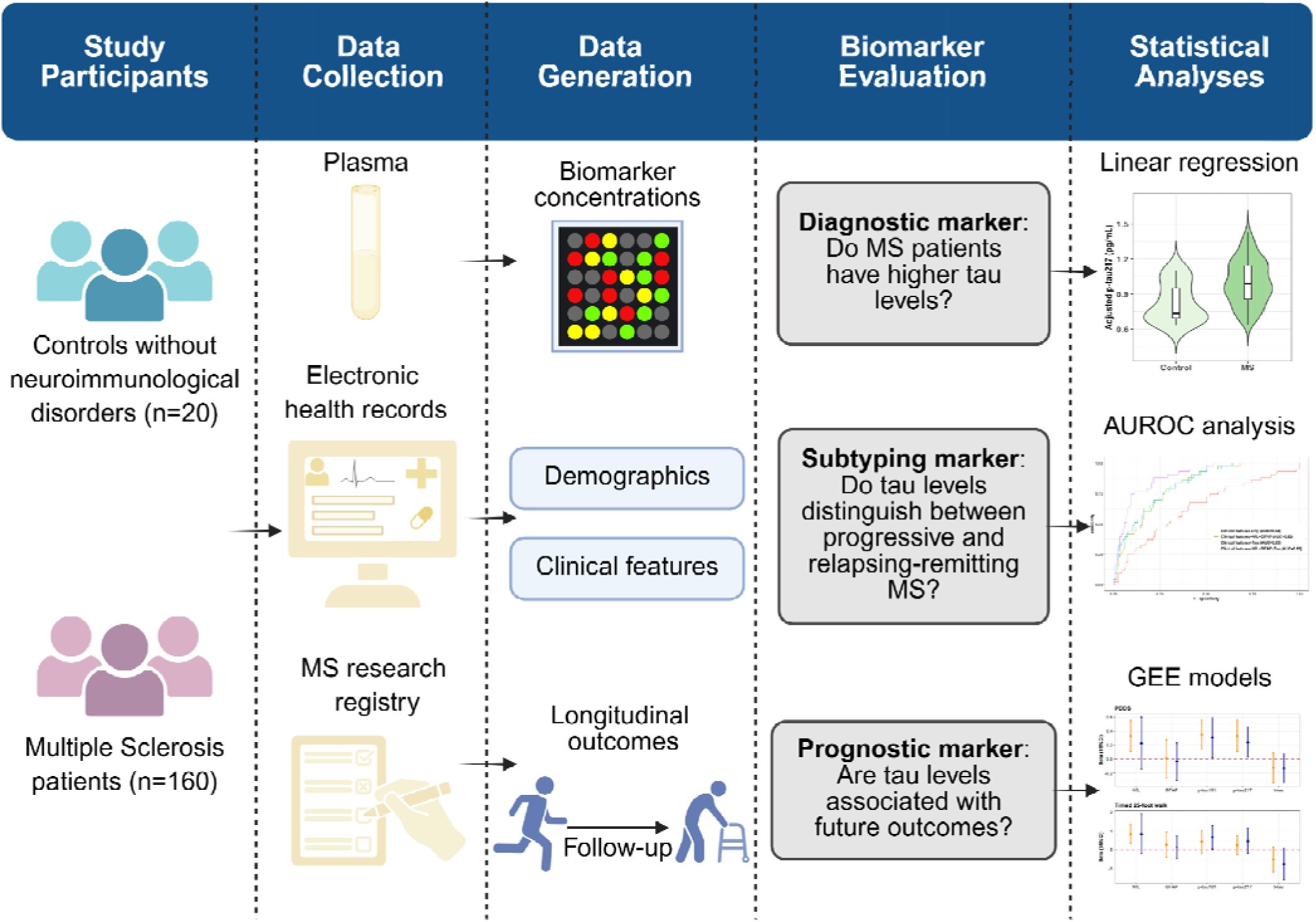
Overall study design. *AUROC*, area under the receiver operating characteristic curve; *GEE*, generalized estimating equat on.

### Exposure: plasma biomarkers

Participants donated venous blood samples during routine clinic visits. Plasma samples were isolated and stored per standard protocol until assay. We selected the first available plasma sample of each participant as baseline. We quantified baseline plasma levels of p-tau181, p-tau217, t-tau, NfL, and GFAP using ultrasensitive single molecule array (Simoa) immunoassays. Detailed assay protocols, quality control procedures, and validation data are in **eMethods-1**.

### Outcomes

Beyond MS diagnosis and subtype, we examined multi-modal patient-reported and rater-assessed disability measures, functional status and imaging outcomes: *e.g.,* Patient Determined Disease Steps (PDDS), age-related MS severity score (ARMSS), timed 25-foot walk (T25-FW), nine-hole peg test (9-HPT), and Symbol Digit Modalities Test (SDMT), retinal nerve fiber layer (RNFL) thickness, and total and gray matter brain volumes. Outcome definitions and procedures are in **eMethods-2**.

### Covariates

We collected demographic and clinical data from cohort registry and confirmed via review of electronic health records: age (at blood sample collection), sex, self-reported race and ethnicity (non-Hispanic white vs. otherwise), height and weight, disease subtypes (RRMS, PPMS, and SPMS), disease duration (years between MS diagnosis and sample collection), DMT effectiveness (none, standard, and high), baseline PDDS (*i.e.,* score reported on the day closest to sample collection), and relapse history one year before the sample collection. Obesity status was based on body mass index (BMI≥30 kg/m^2^) as waist size was unavailable. We combined PPMS and SPMS under progressive MS (PMS), given the modest sample size of each subtype. For DMT, we operationally categorized natalizumab, ocrelizumab, ofatumumab, rituximab and cladribine as high-effectiveness, while dimethyl fumarate, fingolimod, glatiramer acetate, interferon beta, and teriflunomide as standard-effectiveness.

### Statistical analysis

Participant characteristics were presented as mean (SD), median (IQR), or frequencies and proportions, depending on the type and distribution of the data. Using nonparametric methods, we described correlations among biomarkers and biomarker correlation with age, sex, and race and ethnicity. We assessed the association of tau biomarkers with MS diagnosis (*i.e.*, MS vs controls), adjusting for age and sex. We categorized pwMS into RRMS and PMS and tested the difference in tau biomarker values between MS subtypes and to controls. Dunn’s test with Benjamini-Hochberg (BH) corrected p-value was used for pairwise comparison when appropriate.

Among pwMS, we used weighted logistic regression to assess cross-sectional associations between biomarkers and the odds of PMS. Balancing weights were used to account for differences in the distributions of age, sex, race and ethnicity, and disease duration between PMS and RRMS (**eMethods-3**). We first entered the markers separately to compare the individual association of each biomarker (model 1). We then tested the adjusted associations of the biomarkers by entering all biomarkers simultaneously (model 2), to investigate the added value of each marker to other markers. To compare the ability of tau biomarkers to discriminate PMS from RRMS with clinical features (age, sex, race and ethnicity, obesity, disease duration, and DMT effectiveness) and benchmark biomarkers (NfL and GFAP), we used the area under the receiver operating characteristics curve (AUROC) and the decision curve analyses for models with different sets of predictors (*i.e.*, set 1: clinical features alone; set 2: clinical features, NfL and GFAP; set 3: clinical features and tau biomarkers; set 4: all predictors together). AUROC analysis evaluated model accuracy while decision curve analysis assessed prediction utility regarding clinical value (**eMethods-4**). In addition, other standard metrics of model performance including accuracy, F1 score, sensitivity, specificity, positive predictive value (PPV), and negative predictive value (NPV) were estimated with the optimal cut points from 5-fold cross-validation.

Among pwMS with ≥1 assessment of target outcomes taken ≥3 months after completing the baseline blood draw, we examined longitudinal associations between baseline biomarkers and subsequent multi-modal outcomes. We used generalized estimating equation (GEE) models to estimate the relationship between biomarkers with repeatedly measured PDDS, T25-FW, 9-HPT, SDMT, RNFL thickness, normalized total brain volume, and normalized gray matter volume over the follow-up. All GEE models used an unstructured correlation matrix and a robust covariance matrix. For each outcome, we reported the overall and independent change per 1 SD increase in the marker concentration by entering each biomarker separately and then entering all biomarkers simultaneously. All models were adjusted for age, sex, race and ethnicity, disease duration, obesity status, MS subtype, baseline PDDS, DMT effectiveness, and 1-year relapse history. Additionally, we conducted supplementary analyses using tertile-based models and sensitivity checks for outcomes assessed ≥6 months post-baseline. These details are reported in **eMethods-5**.

Statistical significance for the cross-sectional and longitudinal associations was defined as p<0.05. We did not correct for multiple comparisons in these analyses because the goal was to compare tau biomarkers with benchmark GFAP and NfL. Statistical analyses were performed using R version 4.3.2.

### Data sharing statement

Code for analyses and figures is available at https://github.com/xialab2016/tau_biomarker. De-identified data are available upon request to the corresponding author and with permission from the participating institutions.

## Results

### Cohort characteristics

In the clinic-based cohort of 180 participants (pwMS: n=160, 88.9%; controls: n=20, 11.1%) with baseline plasma samples, the predominantly non-Hispanic White (n=164; 91.1%) and women (n=123, 68.3%) demographics are representative of the broader clinic population (**Table 1**). 117 (73.1%) pwMS had RRMS subtype at sample collection. Compared to controls, pwMS had significantly higher levels of plasma p-tau217 (median=1.0 vs 0.7 pg/mL, p=0.04) and NfL (median=14.1 vs 9.0 pg/mL, p<0.01), while other biomarkers had limited diagnostic value in distinguishing pwMS from controls. Correlations among biomarkers (concentrations) and biomarker correlations with age, sex, and race/ethnicity were reported in **Supplementary Results** and **eFigure 1**.

**Table 1.**
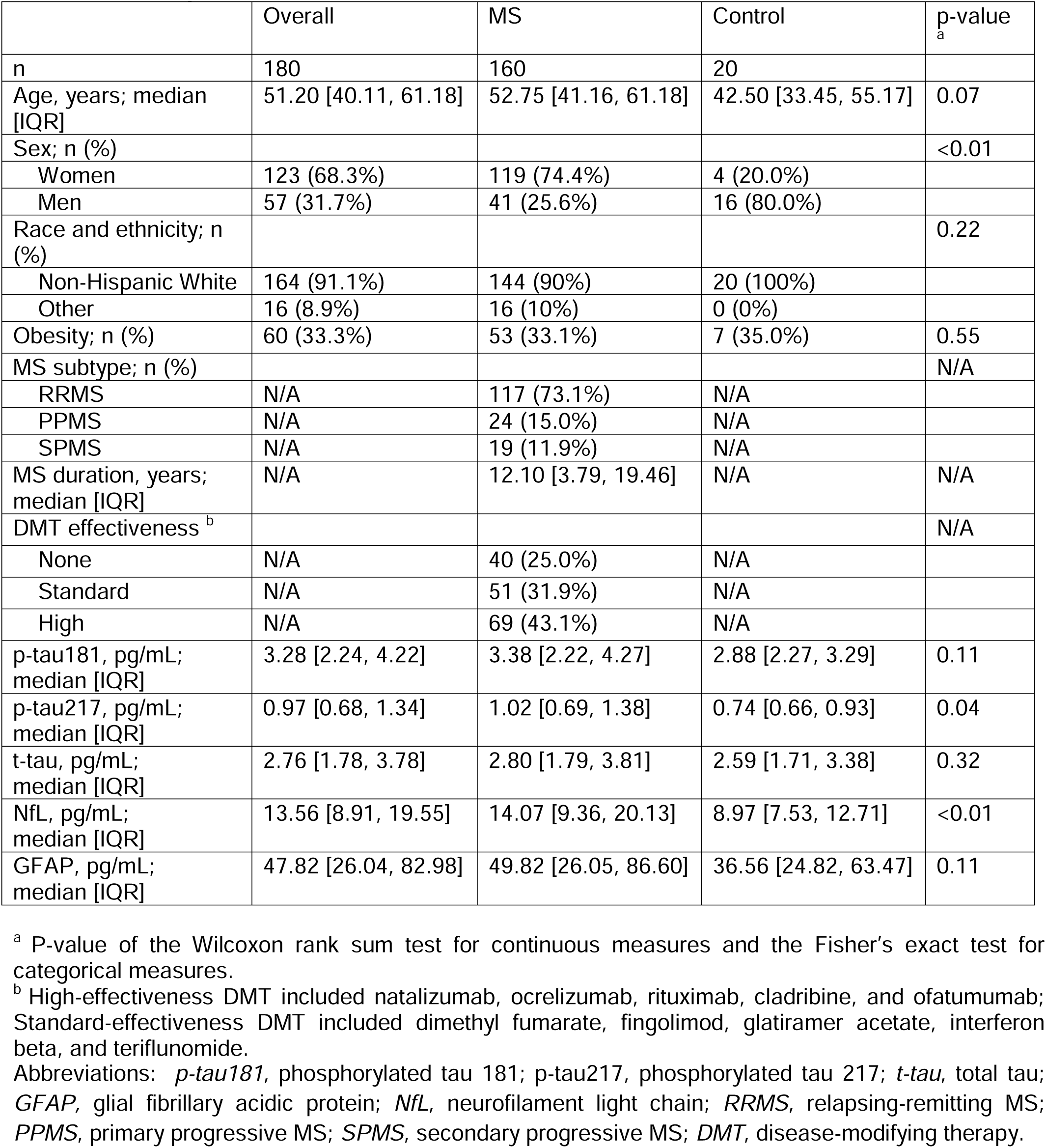
Participant Characteristics.

### Progressive MS and plasma tau biomarkers

Among the 160 pwMS, we applied the balancing weights method to assess the association between biomarker concentrations and the odds of having PMS. The distribution of age, sex, race and ethnicity, and disease duration were comparable between PMS and RRMS after balancing weights (**eTable 1**). In the weighted cohort, elevated p-tau181 (OR [95%CI]=2.30 [1.40,4.13]) and p-tau217 (OR [95%CI]=3.03 [1.75,5.67]) were each associated with higher odds of PMS, whereas t-tau, NfL and GFAP were not (**Figure 2A**). After adjusting for other biomarkers, higher p-tau181(OR [95%CI]=1.36 [1.03,1.80]) and p-tau217 (OR [95%CI]=1.65 [1.04,2.62]) maintained associations with higher odds of PMS (**Figure 2A**).

**Figure 2.**
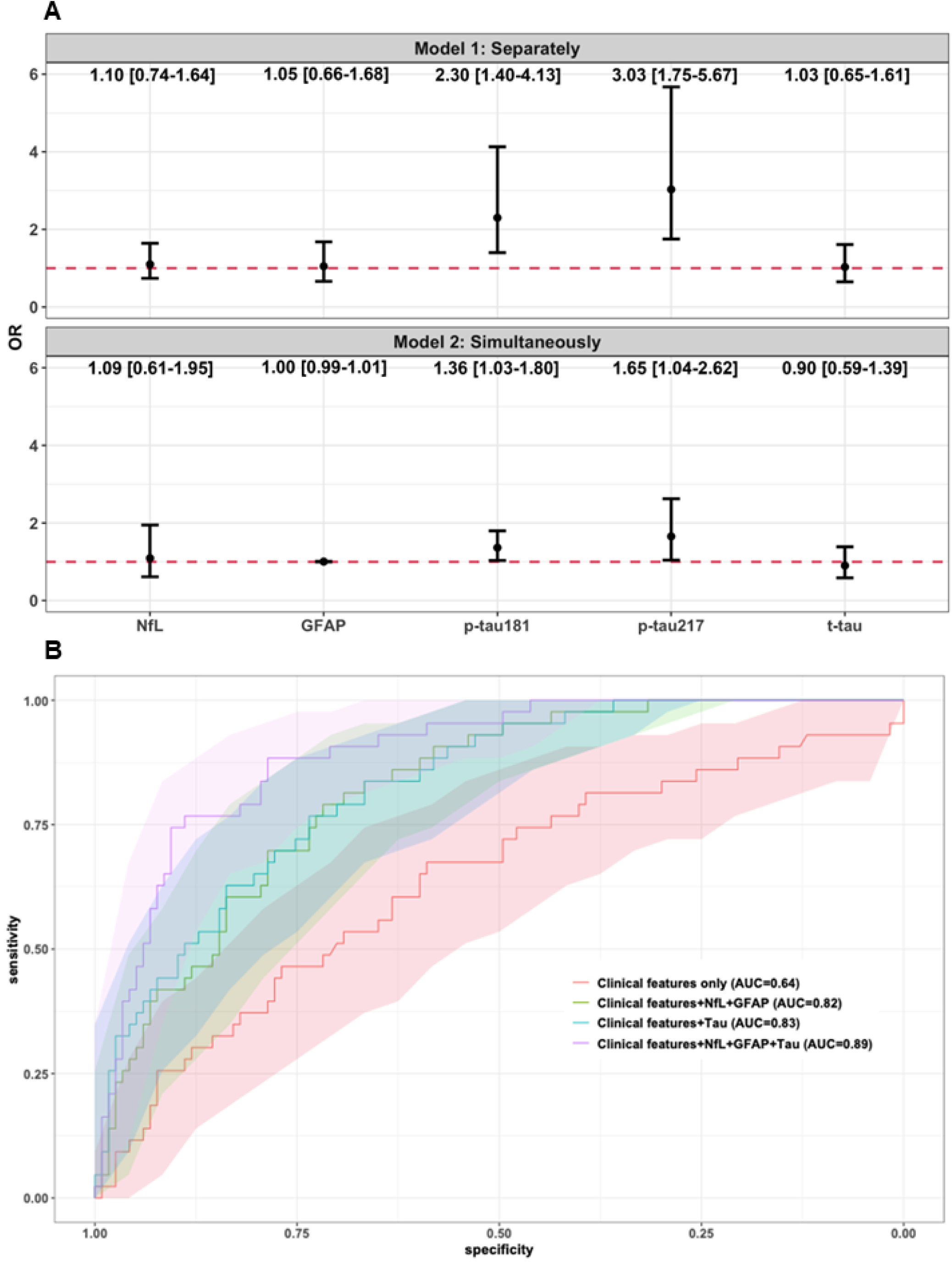
Biomarkers and MS subtypes. **(A) Blood biomarker concentration and odds of progressive MS.** The results were derived from weighted logistic regression models with balancing weights on age, sex, race and ethnicity, and disease duration. Estimates displayed are odds ratio and corresponding 95% CI for a 1 SD increase in each biomarker concentration. The red dashed line represents a null association of OR = 1. In Model 1, separate logistic regression models were run for each biomarker. In Model 2, all markers were entered simultaneously to assess the adjusted association independent of other markers. **(B) Receiver operating curves (ROC) and corresponding 95%CI of different sets of predictors for distinguishing MS subtypes.** The value of area under the curve (AUC) indicates model performance in distinguishing progressive MS from relapse-remitting MS. 50% indicates no difference from chance and 100% indicates a perfect ability with sensitivity and specificity of 100%. Clinical features included age, sex, race and ethnicity, obesity status, disease duration, and DMT effectiveness. **Tau** included p-tau181, p-tau217, and t-tau. Abbreviations: *p-tau181*, phosphorylated tau 181; p-tau217, phosphorylated tau 217; *t-tau*, total tau; *GFAP,* glial fibrillary acidic protein; *NfL*, neurofilament light chain; *OR*, odds ratio.

As predictors, Tau biomarkers (p-tau181, p-tau217, t-tau) along with clinical features, GFAP, and NfL distinguished PMS from RRMS with clinically actionable accuracy (AUC [95%CI]=0.89 [0.84,0.94]; **Figure 2B**; **eTable 2**). This combined feature set outperformed clinical features alone (AUC=0.64 [0.54,0.74]), clinical features with GFAP and NfL (AUC=0.82 [0.75,0.86]), and clinical features with tau biomarkers (AUC=0.83 [0.76,0.89]). Decision curve analysis quantified the clinical utility of models by net benefit. Adding tau biomarkers as predictors to clinical features, NfL and GFAP yielded increased net benefit across a wide range of threshold probability (**eFigure 2**). Considering pwMS with a predicted PMS risk of 0.5 (*i.e.,* at 50% probability threshold, 50% chance to be PMS vs RRMS), incorporation of tau biomarkers added a ∼5% higher net benefit than the prediction model containing clinical features, GFAP and NFL (**eTable 2**). Other metrics of MS subtype classification models are shown in **eTable 3**.

### Multi-modal outcomes and plasma tau markers

The median [IQR] of the cohort follow-up was 3.0 [1.2, 4.7] years. **Figure 3** and **eTable 4** present associations and dose-response trends between baseline biomarkers and longitudinal multi-modal outcomes 3 months post-baseline (*i.e.,* after sample collection), adjusting for age, sex, race and ethnicity, disease duration, obesity status, MS subtype, baseline PDDS, DMT effectiveness, and 1-year relapse history. Results on outcomes 6 months post-baseline are reported in **Supplementary Results**. ***Disability: patient-reported (PDDS) and rater-assessed (normalized age-related MS severity, ARMSS)***

**Figure 3.**
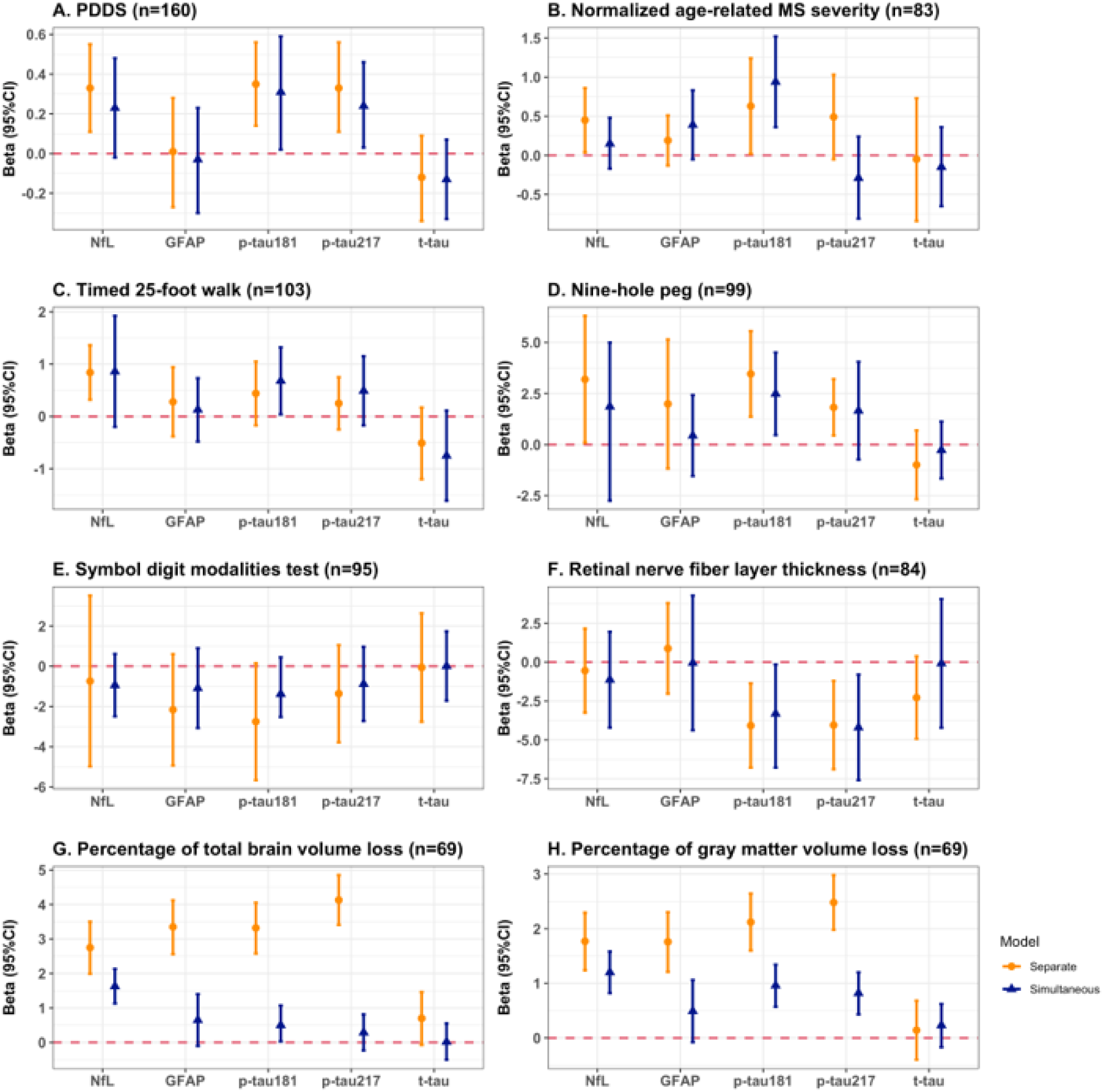
Associations between baseline biomarker concentration and multi-modal outcomes 3 months after baseline. Generalized equation estimation models were used for repeated measured outcomes during the follow-up (**A**, PDDS; **B**, Normalized age-related MS severity; **C**, timed 25-foot walk test; **D**, Nine-hole peg test; **E**, Symbol digit modalities test, correct score; **F**, thickness of retinal nerve fiber layer; **G**, % of total brain volume loss; **H**, % of gray matter volume loss). Estimates and corresponding 95% CI for a 1 SD increase in each marker concentration are displayed. The red dashed line represents a null association of Beta=0. In “separate” models, each biomarker was separately entered. The estimates are the change in the outcome per 1 SD increase in the biomarker value. In the “simultaneous” models, all biomarkers were simultaneously entered. The estimates are the change in the outcome per 1 SD increase in the biomarker value independent of other markers. Abbreviations: *p-tau181*, phosphorylated tau 181; p-tau217, phosphorylated tau 217; *t-tau*, total tau; *GFAP,* glial fibrillary acidic protein; *NfL*, neurofilament light chain; *PDDS,* patient determined disease steps.

All pwMS (n=160) had ≥1 observations of PDDS 3 months post-baseline. Elevated p-tau181, p-tau217 and NfL were each associated with worse PDDS score (beta range:0.33-0.35; **Figure 3A**), and all three biomarkers reached significant dose-response trends when modeling biomarkers as tertiles (**eTable 4**). Estimates are the change in the outcome per 1 SD increase in the biomarker concentration. The association remained significant for p-tau181 (beta [95%CI]=0.31 [0.02,0.59]) and p-tau217 (beta [95%CI]=0.24 [0.03,0.46]), after adjusting for other biomarkers.

83 pwMS had ≥2 observations of EDDS 3 months post-baseline to compute normalized ARMSS. Elevated p-tau181 concentration was associated with worse ARMSS, when modeling separately (beta [95%CI]=0.63 [0.02,1.24]) and simultaneously (beta [95%CI]=0.94 [0.36,1.52]) (**Figure 3B**). We did not observe any significant trend between any plasma biomarker and normalized ARMSS (**eTable 4**).

### Functional outcomes: walk speed (T25FW), manual dexterity (9-HPT), and cognition (SDMT)

When modeling each biomarker individually to predict rater-assessed functional status, elevated p-tau181, p-tau217 and NfL concentrations were associated with worse manual dexterity (higher 9-HPT), while only elevated NfL was associated with slower walk speed (higher T25FW) (**Figure 3C-D**). When modeling all biomarkers simultaneously, 1 SD elevation of p-tau181 was independently associated with worse walk speed (beta [95%CI]=0.68 [0.04,1.32]) and worse manual dexterity (beta [95%CI]=2.49 [0.47,4.50]), whereas no other markers showed significant association. p-tau181 and NfL showed dose-response trends with manual dexterity (**eTable 4**). While both p-tau181 and p-tau217 showed dose-response trends with cognition (lower correct SDMT score) when separating concentrations into tertiles in a data-driven manner, there was no significant association between continuous biomarker concentrations and cognition (**Figure 3E; eTable 4**). Relative to pwMS in the bottom tertile, pwMS in the top tertile of p-tau181 and p-tau217 had 3.15 and 2.41 fewer numbers of SDMT correct scores, respectively.

### OCT outcome: retinal nerve fiber layer (RNFL) thickness

One SD increase in the concentration of p-tau181 and p-tau217 predicted a 4.08µm ([95% CI]= [1.37, 6.78]) and 4.05µm ([95% CI=[1.21, 6.89]) decrease of RNFL thickness during follow-up, respectively (**Figure 3F**). Further, higher baseline p-tau181 (beta [95%CI]=−3.31 [−6.78,−0.16]) and p-tau217 (beta [95%CI]=−4.20 [−7.59,−0.81]) were associated with lower RNFL thickness, independent of other biomarkers, whereas t-tau, NfL, and GFAP were not. A significant trend was detected for p-tau217, as pwMS in the top tertile of p-tau217 had a mean of 7.01µm lower in RNFL thickness relative those in the bottom tertile (**eTable 4**).

### MRI outcome: total brain volume (TBV) and gray matter volume (GMV) normalized to the intracranial volume

All biomarkers except for t-tau were separately associated with normalized TBV and GMV (beta range: 2.75%-4.13% for TBV, 1.76%-2.48% for GMV; **Figure 3G-H)**. After adjusting for other biomarkers, elevated p-tau181 (beta [95%CI]=0.50 [0.03,1.07]) and NfL (beta [95%CI]=1.63 [1.13, 2.13]) were independently associated with greater TBV loss, while elevated p-tau181 (beta [95%CI]=0.96 [0.57, 1.34]), p-tau217 (beta [95%CI]=0.82 [0.43,1.20]), and NfL (beta [95%CI]=1.20 [0.82,1.58]) were independently associated with greater GMV loss. p-tau217 and NfL each showed a significant dose-response trend for total brain and gray matter atrophy (**eTable 4**), while p-tau181, t-tau, and GFAP did not.

## Discussion

In this clinic-cohort study, plasma tau markers showed limited diagnostic value, but potential subtyping and prognosis value in MS. Tau markers did not differentiate pwMS from controls. Higher p-tau181 and p-tau217 levels were associated with increased odds of PMS (vs RRMS), improving subtyping distinction beyond clinical features, NfL, and GFAP. Importantly, plasma p-tau181 and p-tau217 showed potential as prognostic markers as they contributed additional value beyond NfL and GFAP in predicting multi-modal disability, functional, and neuroimaging outcomes.

To our knowledge, this is the first study to comprehensively investigate the clinical potential of *blood*-based tau proteins as MS biomarkers. As an axonal cytoskeletal protein, tau is essential for CNS health and an established biomarker of neurodegeneration (*e.g.,* Alzheimer’s disease).^37,38^ Axonal damage and neuronal loss began in early MS stages.^39,40^ Neuropathological and pre-clinical studies reported abnormally phosphorylated tau in MS that may form neurotoxic tau aggregates.^29,30^ Emerging studies evaluated CSF tau, while few studied blood tau in pwMS.^41^ While detectable, blood tau concentrations are estimated to be ten times lower than CSF.^42^ Recent advances in ultrasensitive immunoassays enable reliable detection of blood-based tau, offering a less invasive and more accessible alternative for biomarker studies. Leveraging ultrasensitive immunoassays, rigorously annotated patient characteristics, and longitudinal multi-modal outcomes, we assessed the associations of plasma tau markers with MS diagnosis, subtype, and prognosis.

First, we reported limited diagnostic value for plasma p-tau biomarkers. Adjusting for age and sex, tau levels in pwMS and controls did not differ significantly. This contrasts with a prior study reporting a lower t-tau concentration in female pwMS than healthy controls, which might be confounded by not adjusting for age (a non-negligible factor) and disease heterogeneity (*i.e.,* duration, subtype, DMT, etc).^43^ After categorizing pwMS into PMS and RRMS here, PMS (but not RRMS) showed significantly higher age- and sex-adjusted levels of p-tau181 and p-tau217 than controls. Consistent with prior studies of CSF p-tau accumulation in PMS, p-tau may be more applicable in progressive forms of MS, whereas NfL seems more relevant in RRMS.^29,30^

Next, we showed the subtyping utility of plasma p-tau biomarkers to differentiate PMS from RRMS. Neurodegeneration largely drives the RRMS transition to SPMS and the worsening disability in PMS.^1,2,44^ As one potential explanation for the observed association between p-tau and PMS, immune cell activation promotes oxidative stress, and subsequent axonal tau phosphorylation and degeneration.^45^ Serum NfL and GFAP could stratify pwMS into different progression and disease activity status.^46^ We observed a similar finding for plasma NfL and GFAP, but crucially highlighted the additive value of plasma p-tau181 and p-tau217 in MS subtyping in combination with NfL and GFAP. The synergistic approach could improve MS subtype classification and inform timely treatment strategies given the growing DMT options for PMS.

Finally, our study suggested the potential prognostic utility of plasma p-tau biomarkers. p-tau181 and p-tau217 (but not t-tau) consistently added predictive value for post-baseline multi-modal outcomes beyond NfL and GFAP. Prior CSF t-tau studies were inconclusive regarding association with MS disability. Marbńez-Yélamos et al. and Virgilio et al. reported CSF t-tau predictive poor outcomes in early RRMS, but Gajofatto *et al.* found CSF t-tau not predictive of disability after ∼6 years of follow-up in pwMS.^47–49^ No study has investigated plasma p-tau and MS progression. We showed consistent p-tau associations with multi-modal patient-reported and rate-assessed clinical outcomes and neuroimaging outcomes. Consistent associations with validated clinical outcomes of disability (*i.e.,* PDDS, normalized ARMSS derived from EDSS) and objective measures of neurodegeneration (*e.g.*, functional performance, RNFL thickness, and brain atrophy) strongly support their prognostic utility.

Our study has several strengths. First, it has the largest sample size to date for investigating fluid (*e.g.,* blood, CSF) tau proteins in pwMS. Second, assays of all biomarkers using the same high-sensitivity platform and standardized protocol enhance the reliability and reproducibility of our exposure assessments. Third, we evaluated the clinical potential of tau biomarkers against benchmark MS biomarkers (*i.e.,* GFAP, NfL) and quantified the added clinical value of tau markers alongside NfL and GFAP. Fourth, we comprehensively examined plasma tau as biomarkers of MS diagnosis, subtype differentiation, and prognosis using rigorous statistical strategies and minimizing confounding biases. Lastly, our clinic-based cohort study incorporated multi-modal longitudinal outcomes collected during routine clinic care, including metrics from clinical OCT and MRI. These clinically available multi-modal outcomes enabled evaluation of plasma tau biomarkers in a real-world setting.

Our study has limitations. The absence of external validation and relatively low proportion of racial and ethnic minorities restrict the broader generalizability of the findings. The pragmatic decision combining PPMS and SPMS as PMS given the sample size limits the biomarker evaluation in these two subtypes, which differ in pathological mechanisms. Future replications in independent cohorts with broader demographic representations and more PMS individuals are crucial. Nevertheless, our study serves as a foundational framework for future investigations of tau markers and tauopathies in MS.

## Conclusion

Plasma tau biomarkers, particularly p-tau181 and p-tau217, are potential biomarkers for MS subtype differentiation and outcome prediction. This study contributes towards the growing research on accessible biomarkers that could supplement existing tools to guide individualized management of pwMS. Beyond further validation of baseline blood tau biomarkers for clinical application, investigation of longitudinal tau biomarkers may provide mechanistic insights into MS progression.

## Abbreviations

P-tau181: phosphorylated tau 181
P-tau217: phosphorylated tau 217
T-tau: total tau
GFAP: glial fibrillary acidic protein
NfL: neurofilament light chain
pwMS: people with multiple sclerosis
MS: multiple sclerosis
ROC: receiver operation curve
AUC: area under the curve
PPV: positive predictive value
NPV: negative predictive value
GEE: generalized estimating equation
PDDS: patient determined disease steps
T25-FW: timed 25-foot walk
9-HPT: nine hole peg test
SDMT: symbol digit modalities test
RNFL: retinal nerve fiber layer
BMI: body mass index
DMT: disease-modifying therapy
CI: confidence interval
SD: standard deviation
PRO: patient-reported outcomes
RRMS: relapsing-remitting MS
PMS: progressive MS
OCT: optical coherence tomography
MRI: magnetic resonance imaging
ARMSS: age-related MS severity

