## Supplemental Materials for "Plasma phosphorylated tau 181 and 217 as biomarkers for multiple sclerosis diagnosis, subtyping, and prognosis"

### **SUPPLEMENTARY MATERIAL**

### **Supplementary Methods**

**1. Plasma collection, Biomarker assays and quality control**

Plasma samples from venous whole blood were isolated within four hours of phlebotomy following standard guidelines, stored at -80°C, and thawed at room temperature before biomarker assay. We centrifuged samples at 4000 x g for 10 minutes to remove particulates. We quantified plasma biomarkers using validated commercially available single molecule array (Simoa) assays from Quanterix on an HD-X instrument (Quanterix, Billerica, MA, USA). NfL and GFAP were measured using the Neurology 2-Plex assay (#103520). P-tau181, p-tau217, and t-tau were measured with the P-tau181 V2 Advantage kit (#103714), the ALZpath Simoa® P-Tau 217 V2 Assay Kit (#104371), and the Tau Advantage Kit (#101552), respectively. We assessed within-run and between-run precision by the coefficient of variation (CV). For each assay, we analyzed samples in three runs. Quality control samples of three different concentrations were analyzed at the beginning and end of each run. The average within-run CVs were 10.6% for NfL, 7.1% for GFAP, 6.1% for p-tau217, 10.9% for p-tau181, and 5.7% for t-tau. The average between-run CVs were 10.6% for NfL, 7.1% for GFAP, 17.3% for p-tau217, 12.6% for p-tau181, and 7.1% for t-tau. We used the average concentration of three runs as the biomarker level.

**2. Outcome assessment procedures and definitions**

For assessing disability, we used a patient-reported outcome based on the PDDS scale and the normalized age-related MS severity score (ARMSS) derived from rater-assessed EDSS observations at ≥2 different time points. Normalized ARMSS is an age-ranked reliable metric that enables longitudinal comparison of disease course among pwMS.^34^ For assessing functional tests, we quantified walking speed, manual dexterity, and cognitive function regularly by the timed 25-foot walk (T25-FW), nine-hole peg (9-HPT), and the symbol digit modalities test (SDMT), respectively. For imaging outcomes, we used clinical data from optical coherence tomography (OCT, ZEISS Cirrus) and 3 tesla brain magnetic resonance imaging (MRI, GE) with NeuroQuant (Cortechs.ai), following clinical quality assurance. In this study, we focused on RNFL thickness, total brain volume, and gray matter volume, which are well-established neuroimaging markers for CNS neurodegeneration.^35,36^ The multi-modal outcomes of this study include: (1) PDDS score; (2) normalized ARMSS; (3) T25-FW: the average of the two completed trials measuring in seconds; (4) 9-HPT: the average of the two completed trials for the dominant hand measuring in seconds; (5) SDMT: the score for the number of correct responses in 90 seconds; (6) RNFL thickness of both eyes measuring in µm; and (7) Total brain and gray matter volume normalized to the intracranial volume.

**3. Balancing weights for confounders adjustment**

We used balancing weights, a statistical approach similar to inverse propensity score weighting, to adjust RRMS individuals to be similar to PMS individuals with respect to demographic factors and achieve causal inference with minimum confounding bias. The balancing weights approach outperforms inverse probability weights with more accurate and stable estimates when the overlap between the exposed and unexposed groups is poor.^50,51^ Our data suggested a great demographic discrepancy between RRMS and PMS. For example, the median [IQR] age of RRMS was 48.1 [36.1, 55.8] years, while 61.1 [56.7, 67.9] years for PMS. Participants were divided into 36 strata based on age category (18-40, 40-60, and 60+ years), sex (male and female), race and ethnicity (non-Hispanic white and otherwise), and disease duration (0-5, 5-15, and 15+ years). All PMS patients were assigned a weight of 1, whereas weights for RRMS patients were calculated as the number of PMS individuals in a particular stratum divided by the number of RRMS individuals. After the application of balancing weights, the distribution of age, sex, race and ethnicity, and disease duration were similar in RRMS and PMS patients (**eTable 1**). Balancing weights were used in logistic regression models to estimate the odds ratios of PMS as compared to RRMS with a 1 SD increase in the biomarker concentration. Second-order polynomial terms for age and disease duration were used to account for residual confounding from the stratification in generating weights.

**4. Decision curve analysis**

Introduced by Vickers and Elkin in 2006, decision curve analysis is a technique to assess the effectiveness of prediction models and diagnostic tests.^52^ This method aims to address the shortcomings of conventional statistical measures like discrimination and calibration, which do not offer direct insights into the clinical usefulness of these tools. Unlike receiver operator characteristic (ROC) analysis, which typically assesses the model accuracy, decision curve analysis assesses the utility of prediction models through the concept of overall net benefit. It’s not uncommon that an accurate model has poor clinical value in terms of net benefit.^53^ In brief, net benefit is calculated across a range of threshold probabilities, defined as the minimum probability of disease/condition at which further intervention would be warranted. Net benefit = sensitivity × prevalence – (1 – specificity) × (1 – prevalence) × w, where w is the odds at the threshold probability. In this use case, the value of the net benefit indicates the benefits of a true positive diagnosis (*e.g.,* identifying a true progressive multiple sclerosis [PMS] patient as PMS) minus the harms of a false positive diagnosis (diagnosing a true relapsing-remitting multiple sclerosis [RRMS] patient as PMS) for any threshold probability. We calculated net benefit over the threshold range (*i.e.*, 0-1) for four models with different sets of predictors (*i.e.*, set 1: clinical features alone; set 2: clinical features, NfL and GFAP; set 3: clinical features and tau biomarkers; set 4: all predictors together). A higher net benefit suggests greater clinical utility of a subtype prediction model at a given threshold probability.

**5. Supplementary analysis for associations between markers and clinical outcomes**

We alternatively modeled the biomarker concentration with categorical tertiles to accommodate dose-response relations, adjusting for the same covariates. Trend analysis was performed by including tertiles as a continuous covariate in models. Based on these models, we reported relative change in each outcome for 2^nd^ and 3^rd^ tertiles relative to the 1^st^ tertile, and p-values for the dose-response trend. In sensitivity analyses, we evaluated whether associations of baseline markers and outcomes 6 months after baseline were consistent with that of 3 months after baseline by restricting included participants being pwMS who had ≥1 assessment of the outcome of interest taken ≥6 months after the baseline blood draw.

### **Supplementary Results**

#### Detailed characteristics of tau markers

We first examined the distributions and correlations of biomarkers in pwMS (**eFigure 1A**). Among the tau-based biomarkers, there were significant positive correlations between p-tau181 and p-tau217 (corr=0.69, p<0.01), and p-tau181 and t-tau (corr=0.27, p<0.01). NfL correlated with GFAP (corr=0.39, p<0.01) and were both positively correlated with p-tau217. We next examined biomarker correlations with age, sex, and race and ethnicity (**eFigure 1B-D**). A nonlinear relation with age was observed for all markers (**eFigure 1B**). T-tau was significantly higher in females than in males (median concentration: 3.13 vs. 2.16 pg/mL; p<0.01; **eFigure 1C**). Otherwise, no significant difference was observed for subgroups of sex and race/ethnicity (**eFigure 1C-D**). **eFigure 1E** demonstrates age and sex-adjusted tau markers across disease diagnosis groups. Tau levels were statistically similar between pwMS and controls, while a significant difference was observed in PMS as compared with controls regarding p-tau181 (adjusted mean: 4.1 vs. 2.8 pg/mL; BH corrected p<0.01) and p-tau217 (adjusted mean: 1.2 vs. 0.8 pg/mL; BH corrected p< 0.01).

#### Detailed multi-modal outcomes and plasma tau markers

The point estimates of associations between baseline markers and subsequent outcomes were qualitatively similar when we applied a longer prediction time window (*i.e.*, 6 vs 3 months post-baseline). While results lost statistical significance for certain associations due to a smaller sample size (*e.g.*, n=160 vs n=142 for the 6- and 3-month post-baseline assessments of PDDS, respectively), general associations remained consistent. For instance, 1 SD increase in concentration of p-tau217 was independently associated with a 0.16 ([95% CI]=[0.02, 0.30]) point increase of PDDS (**eFigure 3; eTable 5**). Likewise, p-tau181 association with normalized ARMSS (beta [95%CI]=0.94 [0.24, 1.64]), walking speed (beta [95%CI]=0.62 [0.07, 1.18]), and manual dexterity (beta [95%CI]=3.00 [0.81, 5.19]) remained significant when modeling biomarkers simultaneously (**eFigure 3; eTable 5**). Finally, 1 SD elevation in p-tau217 concentration was independently associated with worse neuroimaging metrics: 3.66µm thinner RNFL thickness, 0.66% greater loss of TBV, and 0.95% greater loss of GMV **(eTable 5**).

### **Supplementary Figures**

#### **eFigure 1. Correlations among biomarkers and biomarker correlations with age, sex, and race and ethnicity, and disease diagnosis groups.**

**(A) Distribution and correlation of p-tau181, p-tau217, t-tau, NfL and GFAP.** The diagonal is the density probability of each biomarker. The upper diagonal is the Spearman correlation coefficient between each pair. The lower diagonal is the scatter plot for each pair of markers. **(B) Biomarker concentration and age (years)**. Blue lines represent the regression of locally weighted scatterplot smoothing (LOWESS) with 95% CI in gray. **(C) Biomarker concentration and sex. (D) Biomarker concentration and race and ethnicity. (E) Age-sex-adjusted tau values and diagnosis groups.** X-axis represents the diagnosis groups. Y-axis represents age and sex- adjusted values deriving from multivariable linear regression models. Second-order polynomial terms of age were adjusted due to the nonlinearity of biomarker concentration with age. Abbreviations: *p-tau181*, phosphorylated tau 181; p-tau217, phosphorylated tau 217; *t-tau*, total tau; *GFAP,* glial fibrillary acidic protein; *NfL*, neurofilament light chain. Significance thresholds: ‘***’, p<0.001; ‘**’, p<0.01; ‘*’, p<0.05; ‘n.s’, p>0.05.

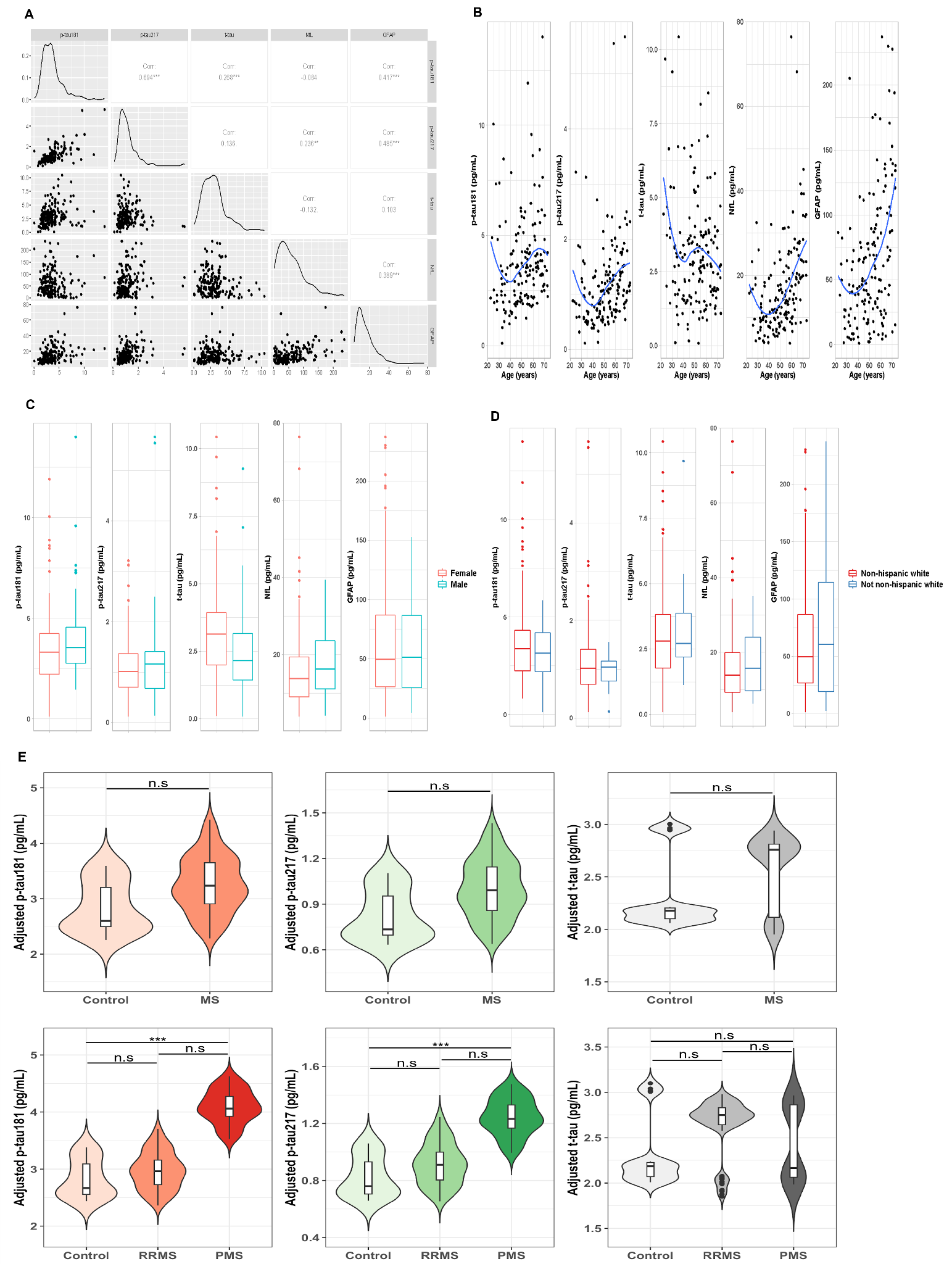

#### **eFigure 2. Decision curves for MS subtype prediction models.**

The value of the net benefit (Y-axis) indicates the benefits of a true positive diagnosis (of MS subtype) minus the harms of a false positive diagnosis for any threshold probability (x-axis). A higher net benefit suggests greater clinical utility of a subtype prediction model at a given threshold probability. For example, at a threshold of 0.5, the probabilities of true PMS among those who were predicted to be PMS by each model (*i.e.*, clinical features alone, clinical features + NfL + GFAP, clinical features + Tau, and clinical features + NfL + GFAP + Tau) are 0%, 4.8%, 5.2%, and 9.9%, respectively. Clinical features included age, sex, race and ethnicity, obesity status, disease duration, and DMT effectiveness. Tau included p-tau181, p-tau217, and t-tau. Abbreviations: *p-tau181*, phosphorylated tau 181; p-tau217, phosphorylated tau 217; *t-tau*, total tau; *GFAP,* glial fibrillary acidic protein; *NfL*, neurofilament light chain.

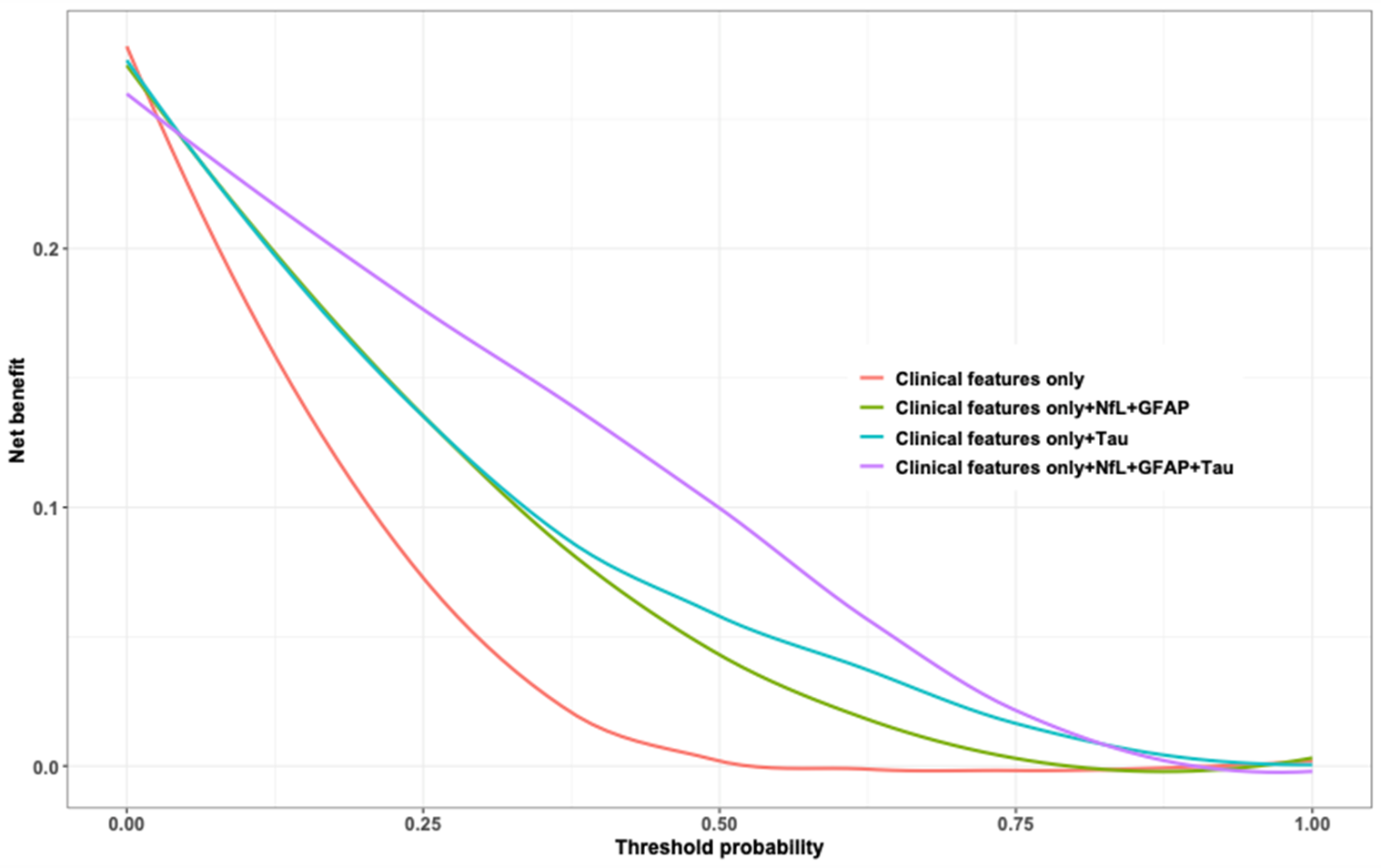

#### **eFigure 3. Associations between baseline biomarker concentration and multi-modal outcomes 6 months after baseline.**

Generalized equation estimation models were used for repeated measured outcomes during the follow-up (**A**, PDDS; **B**, Normalized age-related MS severity; **C**, timed 25-foot walk test; **D**, Nine-hole peg test; **E**, Symbol digit modalities test, correct score; **F**, thickness of retinal nerve fiber layer; **G**, % of total brain volume loss; **H**, % of gray matter volume loss). Estimates and corresponding 95% CI for a 1 SD increase in each marker concentration are displayed. The red dashed line represents a null association of Beta=0. In “separate” models, each biomarker was separately entered. The estimates are the change in the outcome per 1 SD increase in the biomarker value. In the “simultaneous” models, all biomarkers were simultaneously entered. The estimates are the change in the outcome per 1 SD increase in the biomarker value independent of other markers. Abbreviations: *p-tau181*, phosphorylated tau 181; p-tau217, phosphorylated tau 217; *t-tau*, total tau; *GFAP,* glial fibrillary acidic protein; *NfL*, neurofilament light chain; *PDDS,* patient determined disease steps.

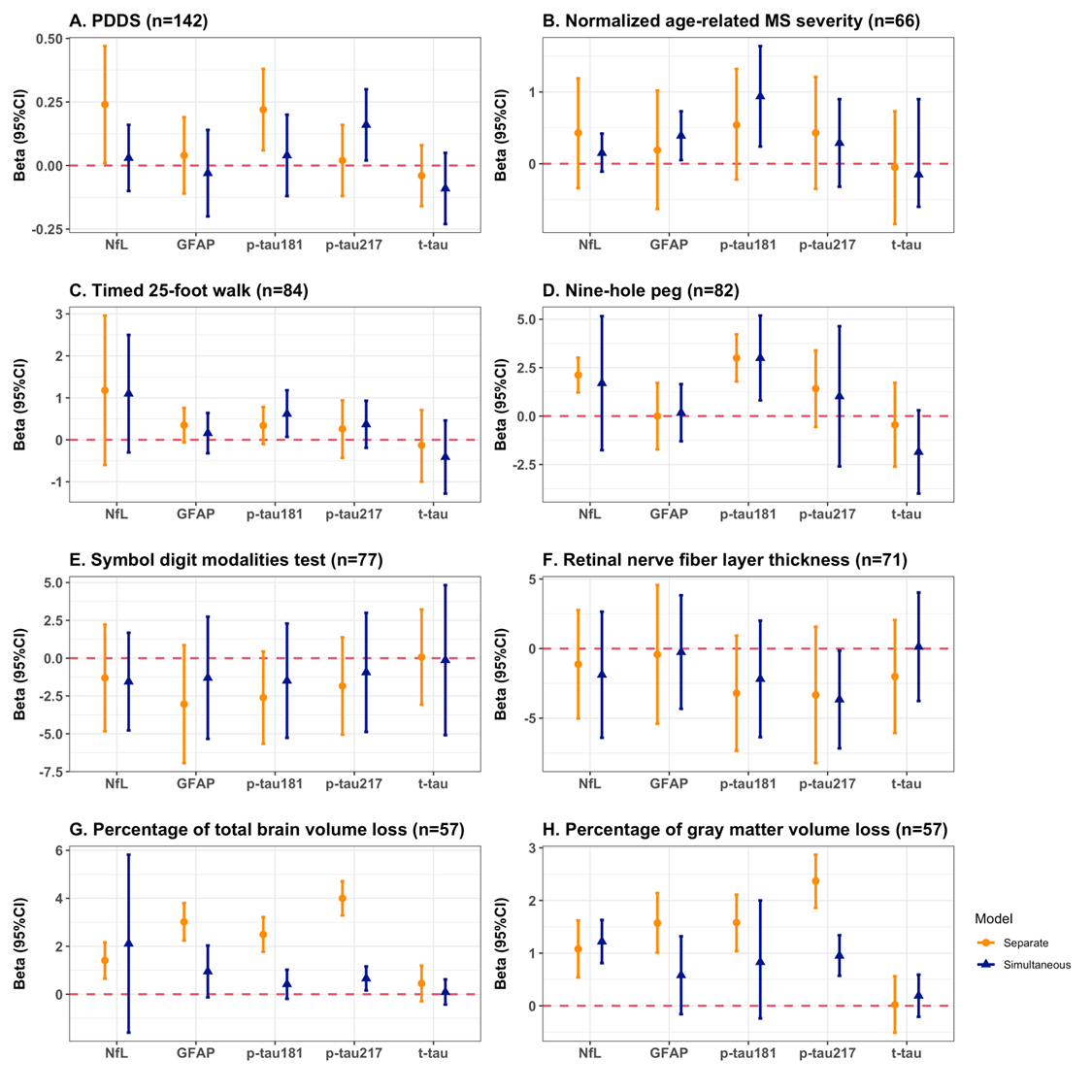

### **Supplementary Tables**

#### **eTable 1. Participant with progressive MS and relapsing-remitting MS with balancing weights**

|  | **Progressive MS** | **Relapsing-remitting MS** | **Relapse-remitting MS with balancing weights** |
| --- | --- | --- | --- |
| n | 43 | 117 | 117 |
| Age, years; median [IQR] | 61.12 [56.66, 67.86] | 48.06 [36.06, 55.82] | 63.84 [57.08, 72.38] |
| Age category; n (%) |  |  |  |
| ≤40 years | 4 (7.1%) | 47 (40.2%) | 10.2 (8.7%) |
| 40-60 years | 15 (35.7%) | 49 (41.9%) | 37.1 (31.8%) |
| >60 years | 24 (57.1%) | 21 (18.0%) | 69.6 (59.5%) |
| Sex; n (%) |  |  |  |
| Women | 21 (47.6%) | 99 (84.6%) | 64.1 (54.8%) |
| Men | 22 (52.4%) | 18 (15.4%) | 52.9 (45.2%) |
| Race and ethnicity; n(%) |  |  |  |
| Non-Hispanic White | 36 (85.7%) | 107 (91.5%) | 100.5 (85.9%) |
| Other | 7 (14.3%) | 10 (8.5%) | 16.5 (14.1%) |
| Disease duration, years; median [IQR] | 15.85 [6.58, 24.91] | 10.43 [3.49, 17.55] | 17.55 [10.88, 22.98] |
| Disease duration category; n (%) |  |  |  |
| ≤5 years | 11 (23.8%) | 38 (32.5%) | 22.7 (19.4%) |
| 5-15 years | 9 (21.4%) | 40 (34.2%) | 21.3 (18.2%) |
| >15 years | 23 (54.7%) | 39 (33.3%) | 73.0 (62.4%) |

#### **eTable 2. Performance for MS subtype prediction models using the area under the curve analysis and the decision curve analysis**

| **Predictors** | **Area under the curve analysis** ^c^ | **Decision curve analysis** | | |
| --- | --- | --- | --- | --- |
|  |  | **Threshold = 0.25** | **Threshold = 0.50** | **Threshold = 0.75** |
| **Set 1**: Clinical features alone ^a^ | 0.64 (0.54, 0.74) | 0.08 | 0 | 0 |
| **Set 2**: Set 1 +NfL+GFAP | 0.82 (0.75, 0.86) | 0.14 | 0.05 | 0.01 |
| **Set 3**: Set 1  +Tau ^b^ | 0.83 (0.76, 0.89) | 0.14 | 0.06 | 0.02 |
| **Set 4**: All | 0.89 (0.84, 0.94) | 0.18 | 0.10 | 0.02 |

Note: Area under the curve analysis evaluated model discrimination as estimated by area under the curve and corresponding 95% CI. Decision curve analysis evaluated the overall clinical utility as estimated by net benefit. See Supplementary Methods. To interpret the decision curve analysis, considering pwMS with a predicted progressive multiple sclerosis (PMS) risk of 0.5 (*i.e.,* at 50% probability threshold, 50% chance to be PMS vs relapsing-remitting multiple sclerosis), incorporation of tau biomarkers added a ~5% higher net benefit than the prediction model containing clinical features, GFAP and NFL (set 2: 0.05; set 4: 0.10).

^a^ Clinical features included age, sex, race and ethnicity, obesity status, disease duration, and DMT effectiveness.

^b^ Tau markers included p-tau181, p-tau217, and t-tau.

^c^ 95%CI from the DeLong test.

Abbreviations: *p-tau181*, phosphorylated tau 181; p-tau217, phosphorylated tau 217; *t-tau*, total tau; *GFAP,* glial fibrillary acidic protein; *NfL*, neurofilament light chain; *DMT*, disease-modifying therapy.

#### **eTable 3. Model metrics of MS subtype classification**

| **Predictors** | **Accuracy** | **F1 score** | **Sensitivity** | **Specificity** | **Positive predictive value** | **Negative predictive value** |
| --- | --- | --- | --- | --- | --- | --- |
| **Set 1**: Clinical features alone ^a^ | 0.53 | 0.77 | 0.25 | 0.95 | 0.52 | 0.73 |
| **Set 2**: Set 1 +NfL+GFAP | 0.74 | 0.81 | 0.35 | 0.79 | 0.73 | 0.84 |
| **Set 3**: Set 1  +Tau ^b^ | 0.70 | 0.81 | 0.38 | 0.85 | 0.78 | 0.82 |
| **Set 4**: All | 0.81 | 0.85 | 0.52 | 0.89 | 0.82 | 0.87 |

Note: Model metrics evaluating the ability to distinguish individuals of progressive MS from relapsing-remitting MS. 5-fold cross-validation with 500 repeats was used to calculate accuracy, F1 score, sensitivity, specificity, positive and negative predictive values.

^a^ Clinical features included age, sex, race and ethnicity, obesity status, disease duration, and DMT effectiveness.

^b^ Tau markers included p-tau181, p-tau217, and t-tau.

Abbreviations: *p-tau181*, phosphorylated tau 181; p-tau217, phosphorylated tau 217; *t-tau*, total tau; *GFAP,* glial fibrillary acidic protein; *NfL*, neurofilament light chain; *DMT*, disease-modifying therapy.

#### **eTable 4. Associations between baseline biomarker concentration and multi-modal outcomes 3 months after baseline**

| Marker | 1 SD increase in marker concentration | | Relative change compared to 1^st^ tertile | | |
| --- | --- | --- | --- | --- | --- |
|  | Separate estimate (95% CI)^A^ | Simultaneous estimate (95% CI)^B^ | 2^nd^ tertile estimate (95% CI)^C^ | 3^rd^ tertile estimate (95% CI)^D^ | Trend p-value^E^ |
|  | **Disability outcome: PDDS (n=160^a^)** | | | | |
| P-tau181 | **0.35 (0.14, 0.56)** | **0.31 (0.02, 0.59)** | 0.27 (-0.35, 0.94) | **0.73 (0.10, 1.36)** | **0.02** |
| P-tau217 | **0.33 (0.11, 0.56)** | **0.24 (0.03, 0.46)** | 0.18 (-0.41, 0.88) | **0.58 (0.05, 1.11)** | **0.01** |
| T-tau | -0.12 (-0.34, 0.09) | -0.13 (-0.33, 0.07) | 0.33 (-0.32, 0.98) | -0.40 (-1.05, 0.24) | 0.23 |
| NfL | **0.33 (0.11, 0.55)** | 0.23 (-0.02, 0.48) | 0.23 (-0.22, 0.68) | **0.77 (0.13, 1.40)** | **0.03** |
| GFAP | 0.01 (-0.27, 0.28) | -0.03 (-0.30, 0.23) | 0.43 (-0.19, 0.94) | 0.20 (-0.41, 0.82) | 0.48 |
|  | **Disability outcome: Normalized age-related MS severity score (n=83^b^)** | | | | |
| P-tau181 | **0.63 (0.02, 1.24)** | **0.94 (0.36, 1.52)** | 0.38 (-0.37, 1.14) | **0.56 (0.38, 0.74)** | 0.09 |
| P-tau217 | 0.49 (-0.05, 1.03) | -0.29 (-0.81, 0.24) | 0.39 (-0.55, 1.32) | 0.47 (-0.18, 1.12) | 0.13 |
| T-tau | -0.05 (-0.84, 0.73) | -0.15 (-0.65, 0.36) | 0.30 (-0.48, 1.08) | 0.50 (-0.29, 1.29) | 0.42 |
| NfL | **0.45 (0.04, 0.86)** | 0.15 (-0.17, 0.48) | 0.36 (-0.96, 1.69) | 0.57 (-0.03, 1.17) | 0.21 |
| GFAP | 0.19 (-0.13, 0.51) | 0.39 (-0.05, 0.83) | 0.38 (-1.58, 2.34) | 0.73 (-0.50, 1.96) | 0.37 |
|  | **Functional outcome: Timed 25-foot walk (n=103^c^)** | | | | |
| P-tau181 | 0.44 (-0.17, 1.05) | **0.68 (0.04, 1.32)** | 0.12 (-1.54, 2.51) | 0.35 (-1.61, 2.31) | 0.13 |
| P-tau217 | 0.25 (-0.25, 0.75) | 0.49 (-0.17, 1.15) | 0.22 (-1.84, 2.28) | 1.90 (-0.31, 4.11) | 0.23 |
| T-tau | -0.51 (-1.20, 0.17) | -0.75 (-1.61, 0.11) | -0.30 (-1.06, 0.46) | 1.71 (-2.33,5.75) | 0.69 |
| NfL | **0.84 (0.32, 1.36)** | 0.86 (-0.20, 1.92) | 0.61 (-1.12, 2.34) | 0.95 (-1.12, 3.02) | 0.18 |
| GFAP | 0.28 (-0.38, 0.94) | 0.13 (-0.48, 0.73) | -0.11 (-1.70, 2.40) | 0.77 (-0.86, 2.39) | 0.33 |
|  | **Functional outcome: Nine-hole peg (n=99^d^)** | | | | |
| P-tau181 | **3.46 (1.37, 5.55)** | **2.49 (0.47, 4.50)** | 2.76 (-0.82, 5.40) | **5.70 (3.26, 8.14)** | **0.01** |
| P-tau217 | **1.82 (0.45, 3.20)** | 1.66 (-0.73, 4.05) | -1.04 (-2.41,0.33) | 2.42 (-2.58, 7.42) | 0.34 |
| T-tau | -0.99 (-2.67, 0.69) | -0.27 (-1.66,1.12) | -0.64 (-5.21, 6.91) | 1.81 (-3.45, 7.07) | 0.44 |
| NfL | **3.19 (0.10, 6.29)** | 1.86 (-2.74, 4.99) | 1.45 (-1.43,4.33) | **4.14 (2.13, 6.15)** | **<0.01** |
| GFAP | 1.99 (-1.17, 5.14) | 0.44 (-1.54, 2.42) | 1.60 (-2.13, 5.62) | 5.44 (-0.26, 11.15) | 0.10 |
|  | **Functional outcome: Symbol digit modalities test (n=95^e^)** | | | | |
| P-tau181 | -2.75 (-5.66, 0.15) | -1.38 (-2.52, 0.44) | -0.24 (-1.81, 1.33) | **-3.15 (-5.13, -1.17)** | **0.03** |
| P-tau217 | -1.36 (-3.78, 1.05) | -0.88 (-2.72, 0.96) | -1.05 (-3.23, 1.13) | **-2.41 (-4.37, -0.45)** | **<0.01** |
| T-tau | -0.06 (-2.76, 2.63) | 0.01 (-1.71, 1.73) | 0.07 (-2.34, 2.48) | -1.13 (-2.95, 0.69) | 0.88 |
| NfL | -0.74 (-4.98, 3.51) | -0.94 (-2.49, 0.61) | -0.05 (-3.96, 3.86) | -1.94 (-5.61, 1.73) | 0.64 |
| GFAP | -2.16 (-4.93, 0.60) | -1.09 (-3.07, 0.89) | -1.35 (-3.11, 0.41) | -3.90 (-7.84, 0.04) | 0.16 |
|  | **OCT outcome: Retinal nerve fiber layer thickness (n=84^f^)** | | | | |
| P-tau181 | **-4.08 (-6.78, -1.37)** | **-3.31 (-6.78, -0.16)** | **-3.15 (-6.10, -0.10)** | **-5.11 (-9.25, -0.97)** | 0.06 |
| P-tau217 | **-4.05 (-6.89, -1.21)** | **-4.20 (-7.59, -0.81)** | -2.83 (-6.55, 0.89) | **-7.01 (-10.60, -3.42)** | **<0.01** |
| T-tau | -2.28 (-4.93, 0.37) | -0.08 (-4.22, 4.06) | -3.06 (-7.20, 1.08) | -0.98 (-4.66, 2.70) | 0.45 |
| NfL | -0.55 (-3.25, 2.15) | -1.13 (-4.21, 1.95) | 0.03 (-4.50, 4.56) | -1.21 (-4.91, 2.49) | 0.83 |
| GFAP | 0.88 (-2.03, 3.79) | -0.05 (-4.38, 4.28) | 1.20 (-2.60, 5.00) | -2.82 (-7.09, 1.45) | 0.44 |
|  | **MRI outcome: Percentage of total brain volume loss (n=69^g^)** | | | | |
| P-tau181 | **3.32 (2.58, 4.05)** | **0.50 (0.03, 1.07)** | 0.78 (-0.10, 1.66) | **0.81 (0.01, 1.61)** | 0.13 |
| P-tau217 | **4.13 (3.41, 4.85)** | 0.29 (-0.23, 0.81) | **1.73 (0.88, 2.58)** | **3.83 (3.04, 4.61)** | **0.03** |
| T-tau | 0.70 (-0.07, 1.46) | 0.02 (-0.50, 0.55) | 0.38 (-0.31, 1.07) | 0.40 (-0.41, 1.21) | 0.88 |
| NfL | **2.75 (1.99, 3.50)** | **1.63 (1.13, 2.13)** | 0.03 (-0.72, 0.78) | **0.94 (0.14, 1.74)** | **<0.01** |
| GFAP | **3.35 (2.56, 4.12)** | 0.65 (-0.10, 1.40) | **1.51 (0.74, 2.29)** | 2.38 (-0.22, 4.98) | 0.21 |
|  | **MRI outcome: Percentage of gray matter volume loss (n=69^h^)** | | | | |
| P-tau181 | **2.12 (1.60, 2.64)** | **0.96 (0.57, 1.34)** | 1.40 (-0.30, 3.11) | **1.60 (0.96, 2.24)** | 0.21 |
| P-tau217 | **2.48 (1.98, 2.98)** | **0.82 (0.43, 1.20)** | 0.25 (-0.38, 0.89) | **0.97 (0.33, 1.60)** | **0.02** |
| T-tau | 0.14 (-0.40, 0.68) | 0.23 (-0.17, 0.62) | 1.50 (-0.41, 3.41) | -0.65 (-1.56, 0.26) | 0.96 |
| NfL | **1.77 (1.24, 2.29)** | **1.20 (0.82, 1.58)** | 0.22 (-0.35, 0.79) | **2.45 (1.85, 3.05)** | **0.04** |
| GFAP | **1.76 (1.21, 2.30)** | 0.49 (-0.08, 1.06) | **0.80 (0.20, 1.40)** | **1.82 (1.02, 2.63)** | 0.11 |

Note: All models were adjusted for age, sex, race and ethnicity, disease duration, obesity status, MS subtype, baseline PDDS, DMT efficacy, and 1-year relapse history.

^A^ Each biomarker was separately entered into the model. The results are the change in the outcome per 1 SD increase in the biomarker concentration.

^B^ All markers were simultaneously entered into the model. The results are the change in the outcome per 1 SD increase in the biomarker independent of other markers.

^C^ The categorical tertiles of each marker were separately entered into the model. The results are the relative change in the outcome for the 2^nd^ tertile relative to the 1^st^ tertile.

^D^ The categorical tertiles of each marker were separately entered into the model. The results are the relative change in the outcome for the 3^rd^ tertile relative to the 1^st^ tertile.

^E^ The tertiles of each marker were separately entered into the model as a continuous variable. P-value for trend is from the Wald test.

^a^ 160 pwMS had ≥1 PDDS measurement after 3 months from the baseline blood draw.

^b^ 83 pwMS had ≥2 EDSS measurement after 3 months from the baseline blood draw.

^c^ 103 pwMS had ≥1 timed 25-foot walk test after 3 months from the baseline blood draw.

^d^ 99 pwMS had ≥1 Nine-Hole peg test after 3 months from the baseline blood draw.

^e^ 95pwMS had ≥1 symbol digit modalities test after 3 months from the baseline blood draw.

^f^ 84 pwMS had ≥1 retinal nerve fiber layer thickness measurement after 3 months from the baseline blood draw.

^g^ 69 pwMS had ≥1 total brain volume measurement after 3 months from the baseline blood draw.

^h^ 69 pwMS had ≥1 gray matter volume measurement after 3 months from the baseline blood draw.

#### **eTable 5. Associations between baseline biomarker concentration and clinical outcomes 6 months after baseline**

| Marker | 1 SD increase in marker concentration | | Relative change compared to 1^st^ tertile | | |
| --- | --- | --- | --- | --- | --- |
|  | Separate estimate (95% CI)^A^ | Simultaneous estimate (95% CI)^B^ | 2^nd^ tertile estimate (95% CI)^C^ | 3^rd^ tertile estimate (95% CI)^D^ | Trend p-value^E^ |
|  | **Disability outcome: PDDS (n=142^a^)** | | | | |
| P-tau181 | **0.22 (0.06, 0.38)** | 0.04 (-0.12, 0.20) | 0.12 (-0.28, 0.62) | 0.25 (-0.10, 0.60) | 0.17 |
| P-tau217 | 0.02 (-0.12, 0.16) | **0.16 (0.02, 0.30)** | 0.26 (-0.11, 0.63) | **0.24 (0.05, 0.43)** | 0.08 |
| T-tau | -0.04 (-0.16, 0.08) | -0.09 (-0.23, 0.05) | 0.11 (-0.47, 0.87) | -0.22 (-0.60, 0.16) | 0.41 |
| NfL | **0.24 (0.01, 0.47)** | 0.03 (-0.10, 0.16) | 0.13 (-0.33, 0.53) | 0.37 (-0.14, 0.88) | 0.29 |
| GFAP | 0.04 (-0.11, 0.19) | -0.03 (-0.20, 0.14) | 0.15 (-0.34, 0.64) | 0.12 (-0.35, 0.59) | 0.60 |
|  | **Disability outcome: Normalized age-related MS severity score (n=66^b^)** | | | | |
| P-tau181 | 0.54 (-0.22, 1.32) | **0.94 (0.24, 1.64)** | 0.28 (-0.17, 0.73) | **0.66 (0.36, 0.96)** | 0.06 |
| P-tau217 | 0.43 (-0.35, 1.21) | 0.29 (-0.32, 0.90) | 0.39 (-0.55, 1.32) | 0.47 (-0.18, 1.12) | 0.17 |
| T-tau | -0.05 (-0.84, 0.73) | -0.15 (-0.60, 0.90) | -0.05 (-0.48, 0.38) | 0.50 (-0.29, 1.29) | 0.60 |
| NfL | 0.43 (-0.34, 1.19) | 0.15 (-0.11, 0.42) | 0.36 (-0.96, 1.69) | **0.67 (0.33, 1.00)** | 0.05 |
| GFAP | 0.19 (-0.63, 1.02) | **0.39 (0.05, 0.73)** | 0.38 (-1.58, 2.34) | 0.73 (-0.50, 1.96) | 0.09 |
|  | **Functional outcome: Timed 25-foot walk (n=84^c^)** | | | | |
| P-tau181 | 0.34 (-0.10, 0.78) | **0.62 (0.07, 1.18)** | 0.14 (-1.87, 2.49) | 0.44 (-1.17, 2.46) | 0.06 |
| P-tau217 | 0.26 (-0.43, 0.94) | 0.37 (-0.19, 0.93) | 0.58 (-1.13, 2.44) | 0.71 (-1.33, 2.75) | 0.36 |
| T-tau | -0.13 (-1.00, 0.71) | -0.41 (-1.28, 0.46) | -0.50 (-1.16, 0.16) | 0.02 (-2.13, 2.17) | 0.49 |
| NfL | 1.18 (-0.60, 2.96) | 1.10 (-0.30, 2.50) | 0.35 (-1.58, 2.35) | 1.93 (-0.08, 2.29) | 0.82 |
| GFAP | 0.35 (-0.06, 0.76) | 0.16 (-0.32, 0.64) | 0.08 (-1.78, 2.42) | 0.05 (-1.77, 2.45) | 0.50 |
|  | **Functional outcome: Nine-hole peg (n=82^d^)** | | | | |
| P-tau181 | **3.00 (1.79, 4.22)** | **3.00 (0.81, 5.19)** | 1.50 (-1.95, 5.85) | **5.36 (1.37, 9.36)** | **<0.01** |
| P-tau217 | 1.42 (-0.56, 3.39) | 1.02 (-2.60, 4.64) | -1.05 (-6.91,4.80) | 2.80 (-3.15,8.75) | 0.80 |
| T-tau | -0.45 (-2.61, 1.72) | -1.85 (-4.00,0.30) | 0.05 (-5.05, 5.15) | 1.71 (-3.00, 6.42) | 0.42 |
| NfL | **2.12 (1.22, 3.02)** | 1.70 (-1.76, 5.16) | 0.17 (-2.76, 3.10) | **6.44 (2.27, 10.62)** | **<0.01** |
| GFAP | 0.00 (-1.72, 1.71) | 0.17 (-1.30, 1.65) | 1.38 (-3.21, 5.96) | 2.20 (-2.59, 6.99) | 0.37 |
|  | **Functional outcome: Symbol digit modalities test (n=77^e^)** | | | | |
| P-tau181 | -2.61 (-5.66, 0.44) | -1.49 (-5.27, 2.29) | -0.35 (-3.82, 3.12) | -3.12 (-7.45, 1.21) | 0.19 |
| P-tau217 | -1.84 (-5.06, 1.38) | -0.94 (-4.88, 3.00) | -0.17 (-4.05, 3.71) | -2.93 (-6.52, 0.66) | 0.07 |
| T-tau | 0.07 (-3.08, 3.22) | -0.13 (-5.09, 4.83) | 0.03 (-4.05, 4.56) | -0.15 (-3.48, 3.18) | 0.87 |
| NfL | -1.31 (-4.84, 2.22) | -1.55 (-4.78, 1.68) | -0.03 (-3.54, 3.48) | -2.19 (-6.48, 2.10) | 0.33 |
| GFAP | -3.04 (-6.94, 0.86) | -1.30 (-5.34, 2.74) | -2.22 (-6.43, 1.99) | -2.98 (-6.88, 0.92) | 0.52 |
|  | **OCT outcome: Retinal nerve fiber layer thickness (n=71^f^)** | | | | |
| P-tau181 | -3.21 (-7.35, 0.93) | -2.18 (-6.37, 2.01) | -2.03 (-5.75, 1.69) | -2.88 (-7.16, 1.18) | 0.58 |
| P-tau217 | -3.33 (-8.23, 1.57) | **-3.66 (-7.17, -0.15)** | -1.15 (-5.13, 2.83) | **-5.03 (-8.70, -1.36)** | **0.03** |
| T-tau | -2.01 (-6.07, 2.05) | 0.13 (-3.77, 4.03) | -2.57 (-6.84, 1.70) | -2.10 (-6.51, 2.31) | 0.92 |
| NfL | -1.13 (-5.03, 2.77) | -1.88 (-6.41, 2.65) | -0.04 (-3.80, 3.72) | -1.93 (-5.79, 1.93) | 0.31 |
| GFAP | -0.42 (-5.40, 4.56) | -0.25 (-4.33, 3.83) | 0.48 (-2.64, 3.60) | -1.07 (-5.17, 3.03) | 0.77 |
|  | **MRI outcome: Percentage of total brain volume loss (n=57^g^)** | | | | |
| P-tau181 | **2.49 (1.77, 3.22)** | 0.42 (-0.19, 1.02) | 0.36 (-0.41, 1.14) | 0.72 (-0.09, 1.52) | 0.19 |
| P-tau217 | **4.00 (3.29, 4.71)** | **0.66 (0.16, 1.16)** | **2.45 (1.60, 3.31)** | 3.38 (-1.21, 7.07) | 0.24 |
| T-tau | 0.45 (-0.29, 1.19) | 0.09 (-0.43, 0.62) | 0.28 (-0.33, 0.89) | 0.38 (-0.47, 1.22) | 0.52 |
| NfL | **1.41 (0.65, 2.16)** | 2.11 (-1.60, 5.82) | 0.23 (-0.49, 0.94) | **1.64 (0.83, 2.46)** | **<0.01** |
| GFAP | **3.02 (2.24, 3.80)** | 0.95 (-0.13, 2.03) | **1.56 (0.78, 2.35)** | 3.19 (-0.09, 6.48) | 0.27 |
|  | **MRI outcome: Percentage of gray matter volume loss (n=57^h^)** | | | | |
| P-tau181 | **1.58 (1.04, 2.11)** | 0.83 (-0.24, 2.00) | 0.12 (-0.30, 0.54) | **1.03 (0.33, 1.74)** | 0.17 |
| P-tau217 | **2.37 (1.86, 2.87)** | **0.95 (0.57, 1.34)** | 0.66 (-0.01, 1.33) | **1.70 (0.07, 3.33)** | **0.04** |
| T-tau | 0.02 (-0.51, 0.56) | 0.19 (-0.21, 0.59) | 0.67 (-0.06, 1.40) | -0.04 (-1.99, 1.91) | 0.99 |
| NfL | **1.08 (0.54, 1.62)** | **1.22 (0.81, 1.63)** | 0.23 (-0.32, 0.79) | **2.92 (2.34, 3.49)** | 0.07 |
| GFAP | **1.57 (1.01, 2.14)** | 0.58 (-0.16, 1.32) | **0.68 (0.08, 1.28)** | 0.69 (-0.07, 1.45) | 0.45 |

Note: All models were adjusted for age, sex, race and ethnicity, disease duration, obesity status, MS subtype, baseline PDDS, DMT efficacy, and 1-year relapse history.

^A^ Each biomarker was separately entered into the model. The results are the change in the outcome per 1 SD increase in the biomarker concentration.

^B^ All markers were simultaneously entered into the model. The results are the change in the outcome per 1 SD increase in the biomarker independent of other markers.

^C^ The categorical tertiles of each marker were separately entered into the model. The results are the relative change in the outcome for the 2^nd^ tertile relative to the 1^st^ tertile.

^D^ The categorical tertiles of each marker were separately entered into the model. The results are the relative change in the outcome for the 3^rd^ tertile relative to the 1^st^ tertile.

^E^ The tertiles of each marker were separately entered into the model as a continuous variable. P-value for trend is from the Wald test.

^a^ 142 pwMS had ≥1 PDDS measurement after 3 months from the baseline blood draw.

^b^ 66 pwMS had ≥2 EDSS measurement after 3 months from the baseline blood draw.

^c^ 84 pwMS had ≥1 timed 25-foot walk test after 3 months from the baseline blood draw.

^d^ 82 pwMS had ≥1 Nine-Hole peg test after 3 months from the baseline blood draw.

^e^ 77 pwMS had ≥1 symbol digit modalities test after 3 months from the baseline blood draw.

^f^ 71 pwMS had ≥1 retinal nerve fiber layer thickness measurement after 3 months from the baseline blood draw.

^g^ 57 pwMS had ≥1 total brain volume measurement after 3 months from the baseline blood draw.

^h^ 57 pwMS had ≥1 gray matter volume measurement after 3 months from the baseline blood draw.
